# The impact of vaccination frequency on COVID-19 public health outcomes: A model-based analysis

**DOI:** 10.1101/2023.01.26.23285076

**Authors:** Madison Stoddard, Lin Yuan, Sharanya Sarkar, Debra van Egeren, Shruthi Mangalaganesh, Ryan P. Nolan, Michael S. Rogers, Greg Hather, Laura F. White, Arijit Chakravarty

## Abstract

While the rapid deployment of SARS-CoV-2 vaccines had a significant impact on the ongoing COVID-19 pandemic, rapid viral immune evasion and waning neutralizing antibody titers have degraded vaccine efficacy. Nevertheless, vaccine manufacturers and public health authorities have a number of levers at their disposal to maximize the benefits of vaccination. Here, we use an agent-based modeling framework coupled with the outputs of a population pharmacokinetic model to examine the impact of boosting frequency and durability of vaccinal response on vaccine efficacy. Our work suggests that repeated dosing at frequent intervals (multiple times a year) may offset the degradation of vaccine efficacy, preserving their utility in managing the ongoing pandemic. Our work relies on assumptions about antibody accumulation and the tolerability of repeated vaccine doses. Given the practical significance of potential improvements in vaccinal utility, clinical research to better understand the effects of repeated vaccination would be highly impactful. These findings are particularly relevant as public health authorities worldwide seek to reduce the frequency of boosters to once a year or less. Our work suggests practical recommendations for vaccine manufacturers and public health authorities and draws attention to the possibility that better outcomes for SARS-CoV-2 public health remain within reach.

## Introduction

As the ongoing COVID-19 pandemic approaches its fourth year, the utility of vaccines in mitigating the death and disability burden of SARS-CoV-2 continues to evolve. Initial reports were consistent with strong vaccinal protection against symptomatic disease [1–3], giving rise to the hope that vaccines could be used to achieve herd immunity to SARS-CoV-2. However, this promise was quickly undermined by rapid declines in vaccinal efficacy against infection (VE_i_) [4,5] driven by waning antibody titers [6–9] and viral immune evasion [9–13].

With herd immunity off the table, public health organizations pivoted to relying on vaccinations to manage the mortality burden of COVID-19, even as spread continued. As vaccinal efficacy against severe disease (VE_s_) was initially very high [1–3], this strategy contributed to a lowering of the infection fatality rate for SARS-CoV-2 [14]. Unfortunately, continued viral evolution has degraded VE_s_ [15–18], although it is partially restored with updated boosters [19–21] targeting newer variants.

Despite their limitations, the current crop of SARS-CoV-2 vaccines continues to form the centerpiece of public health strategies to manage the death and disability burden of COVID-19. At present, there are few nonpharmaceutical interventions (NPIs) mandated in any setting [22–24], while options for treatment of serious disease are limited [25,26] and in some cases (the monoclonal antibodies) have been rendered obsolete by viral evolution [27–29].

On the bright side, immunological correlates of protection for SARS-CoV-2 have been established, which is a boon to rational optimization of vaccine performance. Neutralizing antibody (nAb) titers are a validated correlate of immune protection [30–32] for SARS-CoV-2. nAb titers normalized to mean convalescent titer (from the same study) have been shown to fit well to a nonlinear dose-response relationship that is predictive of reported vaccinal protection across a range of different vaccines [33]. Two such dose-response curves exist, one linking nAb titers to protection against symptomatic infection, and one linking nAb titers to protection against severe COVID-19 outcomes. These relationships have held up across a range of studies [34,35] and against newly emerging variants [21,36–39]. In these studies, nAb titers have been demonstrated to predict waning VE_i_ due to pharmacokinetic effects, as well as due to viral immune evasion. Waning nAb titers have also been demonstrated to be predictive of loss of VE_s_. It bears mentioning that the observed reductions in VE_s_ are inconsistent with the widely held perception [40–42] that the observed durability of T-cell responses [43–45] would provide sustained vaccinal protection against severe disease. (See Supplementary Section S1 for further discussion on the role of T-cells in the vaccinal and natural immune response to SARS-CoV-2).

At present, the utility of vaccines in managing the mortality and morbidity burden of COVID-19 is limited and is facing pressure from a number of different angles (antibody waning, viral evolution and lack of uptake). A recent CDC study showed that the vaccine effectiveness (a measure of VE_i_) of a bivalent mRNA COVID-19 booster received after 2 or more doses of monovalent vaccines ranged from 43% (for the 18-49 age group) to 22% (for the over-65 age group) [20]. When it comes to severe acute disease, VE_s_ for a newly boosted individual is now 56% [46], a steep decline from the originally reported VE_s_ (∼100%) for COVID-19 mRNA vaccines [2,47]. (However, an increase in immunity due to infection among the unvaccinated may account for some of this apparent reduction in vaccine effectiveness [48]). These observed losses in VE_s_ and VE_i_ contrast with the positions of public health organizations, such as the CDC and WHO, who have stated their goal to move to an annual or less frequent boosting schedule for SARS-CoV-2 [49,50]. For the US population, this would represent a sharp (about three-fold) reduction in recommended vaccine dosing frequency relative to the current pace.

In addition to population-level waning in nAb titers, significant inter-individual variation in the strength and durability of the nAb response also complicates the picture. In our prior work, we applied mixed-effects modeling to published SARS-CoV-2 nAb titers post-vaccination [51] and found a wide range of half-lives, with a 95% confidence interval ranging from 33-320 days. Lastly, as the pandemic has progressed, it has become clear that the delayed post-acute sequelae of COVID-19 (“Long COVID”) also represent an important component of the morbidity burden of SARS-CoV-2 infections [52,53], with the potential for substantial impact on population-level health and economic outcomes [54–56]. The risk of long COVID upon infection has also been shown to be only modestly reduced by vaccination [53,57,58].

Given our reliance on vaccines as a COVID-19 control measure and the observed decline in vaccine efficacy, creating a strong quantitative framework for understanding vaccine performance is helpful from a practical standpoint. At this juncture in the pandemic, it may be particularly helpful to ask two questions about SARS-CoV-2 vaccines: “are there ways to use the existing vaccines more effectively through optimization of boosting frequency?” and “what should vaccine makers focus on for the next generation of vaccines?” To answer these questions, we developed a longitudinal PK/PD model of nAb kinetics and coupled it with an agent-based modeling framework. Our model combines population heterogeneity in the durability of the nAb response with the dose-response relationships linking nAb titers to protection from mild and severe disease. We have used this modeling framework to examine population heterogeneity in vaccinal protection over time and in response to viral immune evasion. We formulate and test a potential strategy for improving the practical utility of existing vaccines by altering the dosing interval of the vaccines. We also extend our investigation to hypothetical vaccines with improved durability of response.

## Results

### Boosting frequency determines vaccine efficacy throughout the population

We simulated SARS-CoV-2 spread under endemic conditions to evaluate the relationship between booster frequency and the range of outcomes in the boosted population. The schematic in Figure 1 shows the general structure of the model. The model simulates outcomes for 100,000 individuals over 10 years in an agent-based framework. Each individual is assigned a set of fixed properties (including nAb half-lives, age, contact rate, and vaccination status), and nAb titers are updated at each individual’s vaccine and infection. At each model time step, the force of infection (the product of the number of active infections and the intrinsic reproductive number R_0_ divided by the duration of infection) determines how many individuals in the population are drawn for exposure. Individuals are randomly drawn for exposure with a likelihood proportional to their individual contact rates. Exposed individuals stochastically become infected according to their risk of infection upon exposure, which is determined based on their total nAb titer. A successful infection boosts the individual’s nAb titers by a fixed multiple and increases the count of active infections. Vaccination occurs at a fixed frequency and boosts vaccinated individuals’ vaccine nAb titers.

**Figure 1:**
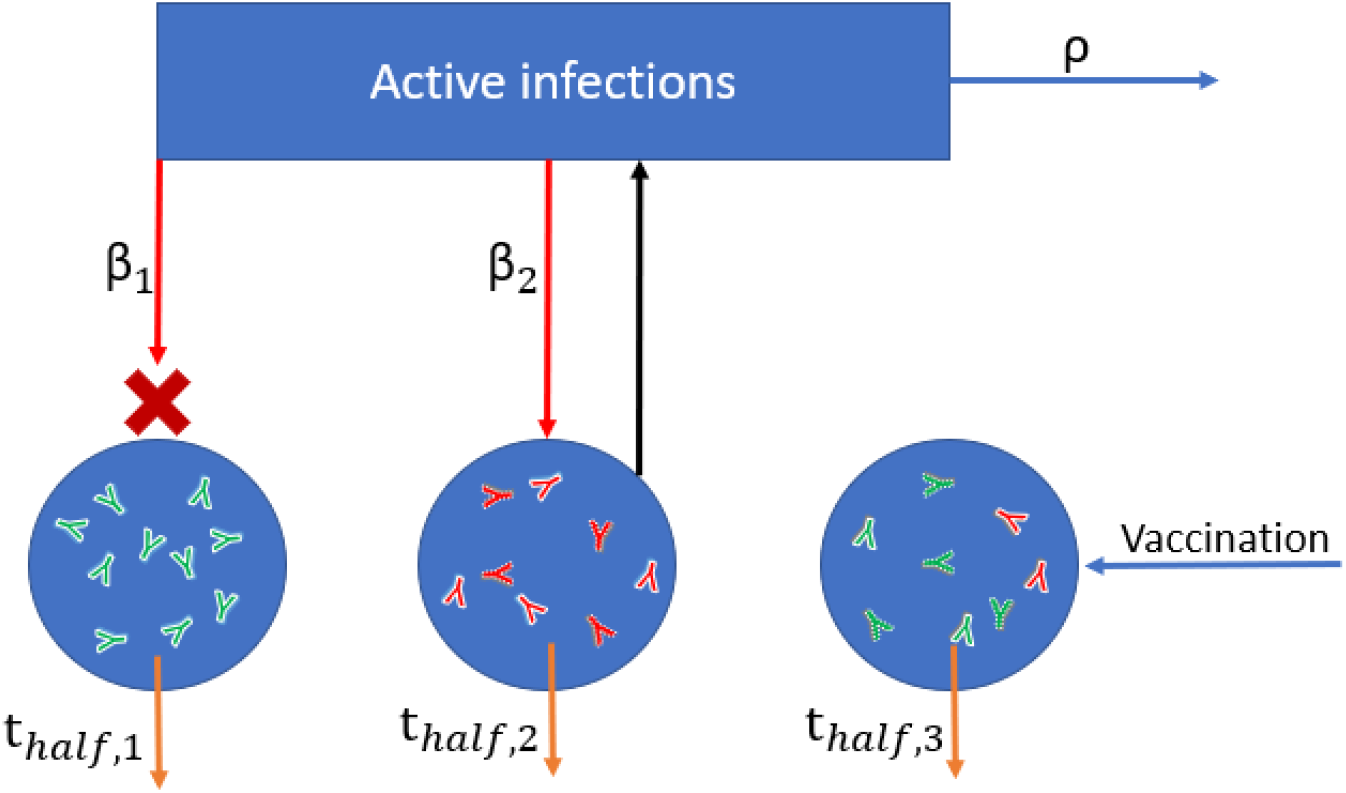
Schematic of simplified agent-based model used to simulate probabilities of infection and adverse outcomes based on nAb titers. The model simulates exposures (red arrows) according to individual contact rates (β_1_, β_2_) and the force of infection (active infections multiplied by R_0_). Exposure succeeds or fails to induce infection probabilistically according to the level of protection afforded by an individual’s combined infection and vaccine nAb titer. nAbs decay over time according to each individual’s half-life (t_half_), and nAb boosting occurs through vaccination (green antibodies) and infection (red antibodies). The number of active infections is tracked, increasing by one for each successful infection and decaying according to the recovery rate (ρ).

We assumed that 50% of the population receives regular boosters, which is roughly consistent with US first booster uptake [59]. The distributions of outcomes in both unvaccinated and vaccinated subpopulations are provided. We demonstrate that once-annual boosting provides some benefit for reducing frequency of infection and risk of death among the vaccinated (Figures 2A-C). In a given year, over 90% of vaccinated individuals are expected to experience SARS-CoV-2 infections. On average, the median boosted individual experiences approximately 1.5 infections per year, while the unvaccinated median individual is infected slightly more than twice yearly. The risk of death for the median unvaccinated individual is predicted to be approximately half that of the median unvaccinated individual.

**Figure 2:**
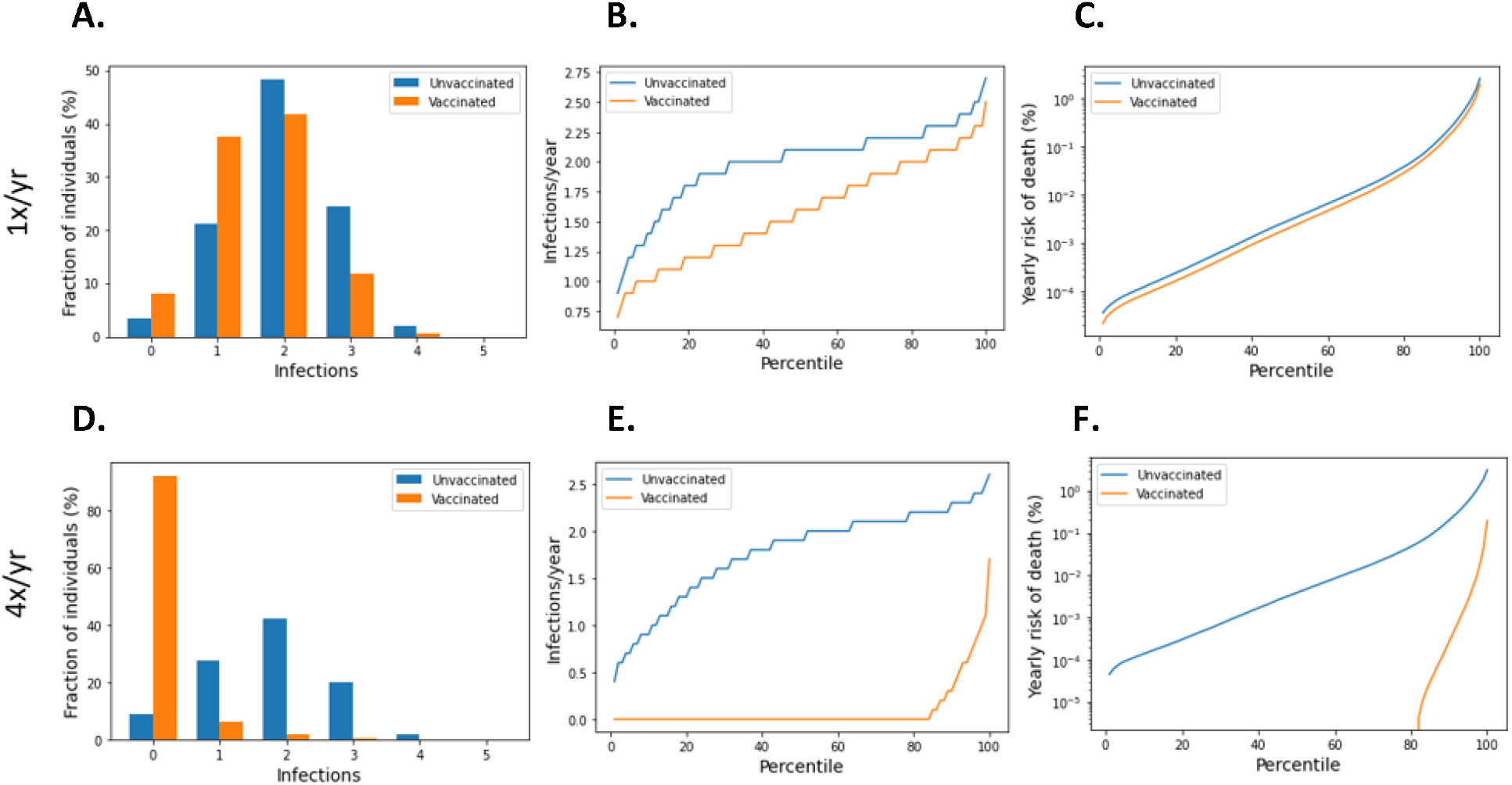
Frequency of boosting determines frequency of infection and risk of COVID-19 death among the vaccinated. On a short-term basis, variation in infection risk is driven by interindividual heterogeneity in biology and behavior as well as stochasticity. In the long-term, interindividual heterogeneity dominates stochasticity in driving individual infection frequency and severe disease risk. For a once-yearly booster frequency, **A**. the distribution of infection counts in a single year, **B**. distribution of individual infection frequencies over a 10-year simulation, and **C**. interindividual heterogeneity in yearly risk of COVID-19 death. For a four-times yearly booster frequency, **D**. the distribution of infection counts in a single year, **E**. distribution of individual infection frequencies over a 10-year simulation, and **F**. interindividual heterogeneity in yearly risk of COVID-19 death.

The outcomes for the vaccinated population improve as boosting frequency improves (Figures 2D-F, Supplementary Figures S5-S6). The vast majority of individuals receiving four boosters per year are not infected in any given year (Figure 2D)-over 80% of these individuals are never infected within a 10-year simulation period (Figure 2E). Annual risk of death greater than 0.1% is extremely rare in the quarterly-boosted population (<2%), while 15% of the unvaccinated population experiences a risk of this magnitude (Figure 2F).

Breakthrough infections under frequent boosting schedules are driven by poor nAb kinetics Although most individuals avoid SARS-CoV-2 infection entirely under a quarterly boosting schedule, 17% of boosted individuals are expected to become infected at any frequency over a 10-year period, with 10% experiencing infection at least every other year (Figure 2E). In Figure 3A, we demonstrate that infection despite quarterly revaccination is strongly predicted by short vaccine nAb half-life. While many individuals with vaccine nAb half-lives less than 50 days are infected, no one with a half-life greater than 50 days is infected in a 10-year simulation. These infections impose significant risk of death, especially for those with the least persistent vaccine antibodies (Figure 3B). However, identification of these individuals and application of more frequent boosting could improve outcomes in this population. We found that six vaccinations per year could suppress infection in those with the shortest vaccine nAb half-lives observed in the immunocompetent population (Figure S2).

**Figure 3:**
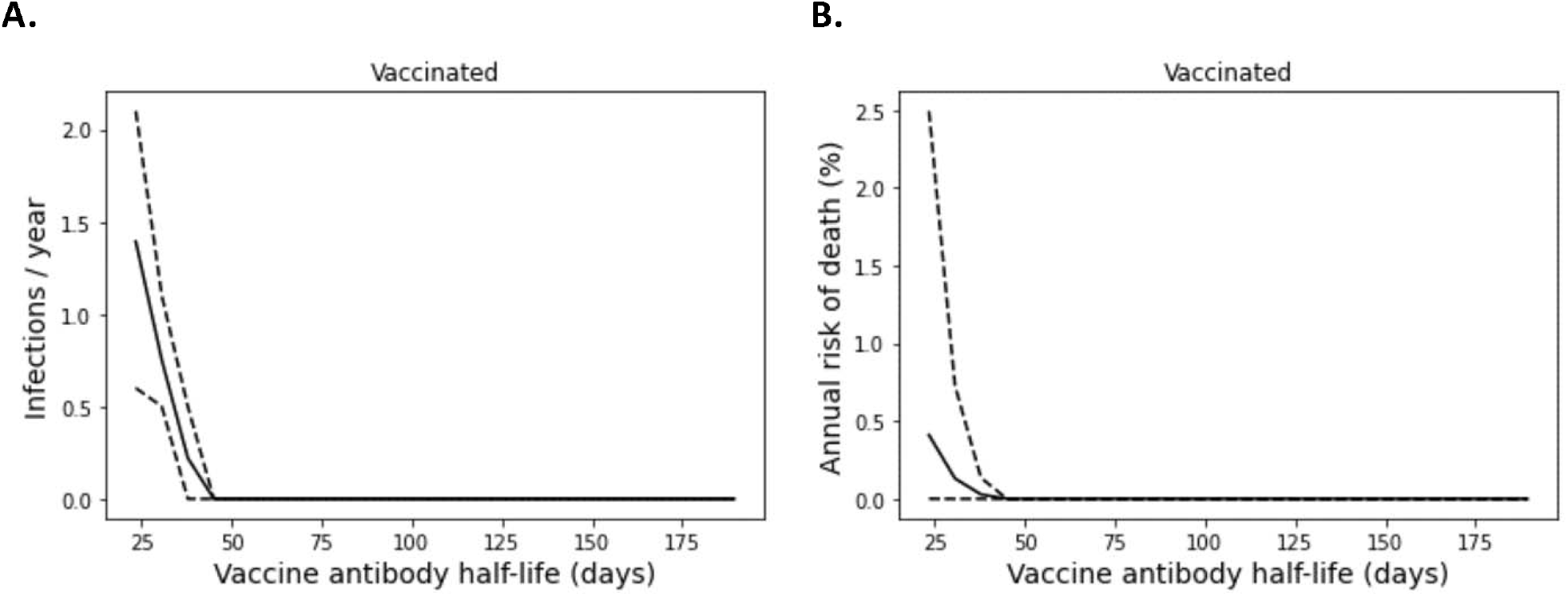
Infection despite four times yearly vaccination is strongly predicted by short vaccine antibody half-lives. **A**. Average infection frequency and **B**. average yearly risk of death over a 10-year simulation. Dashed lines represent the 90% population interval.

### High compliance with frequent boosting could suppress omicron spread

Figure 2 addresses the distribution of individual outcomes under various boosting regimes, while Figure 4 shows the population-level impact of boosting frequency and compliance. Frequent boosting coupled with high compliance is predicted to substantially reduce the impact of COVID-19 at the population level. Despite the high transmissibility of omicron (R_0_ = 8.2 [60–62]), complete suppression of spread is possible with a high degree of compliance and frequent boosting (i.e. approximately 90% compliance with boosters every three months, or perfect compliance with boosters every four months) (Figure 4A). The vaccine’s impact on yearly death tolls is even more dramatic (Figure 4B). In the absence of complete suppression of SARS-CoV-2 spread, increased vaccination coverage and frequency can reduce yearly death tolls. For example, if 50% of the population is vaccinated, an increase in vaccination frequency from once yearly to twice yearly could avert approximately 40,000 US COVID-19 deaths. Increasing vaccination coverage to 90% could prevent an additional 50,000 US COVID-19 deaths. As shown in Figure S3, vaccines with superior kinetics could further reduce infections and deaths in addition to widening the space for complete disease suppression.

**Figure 4:**
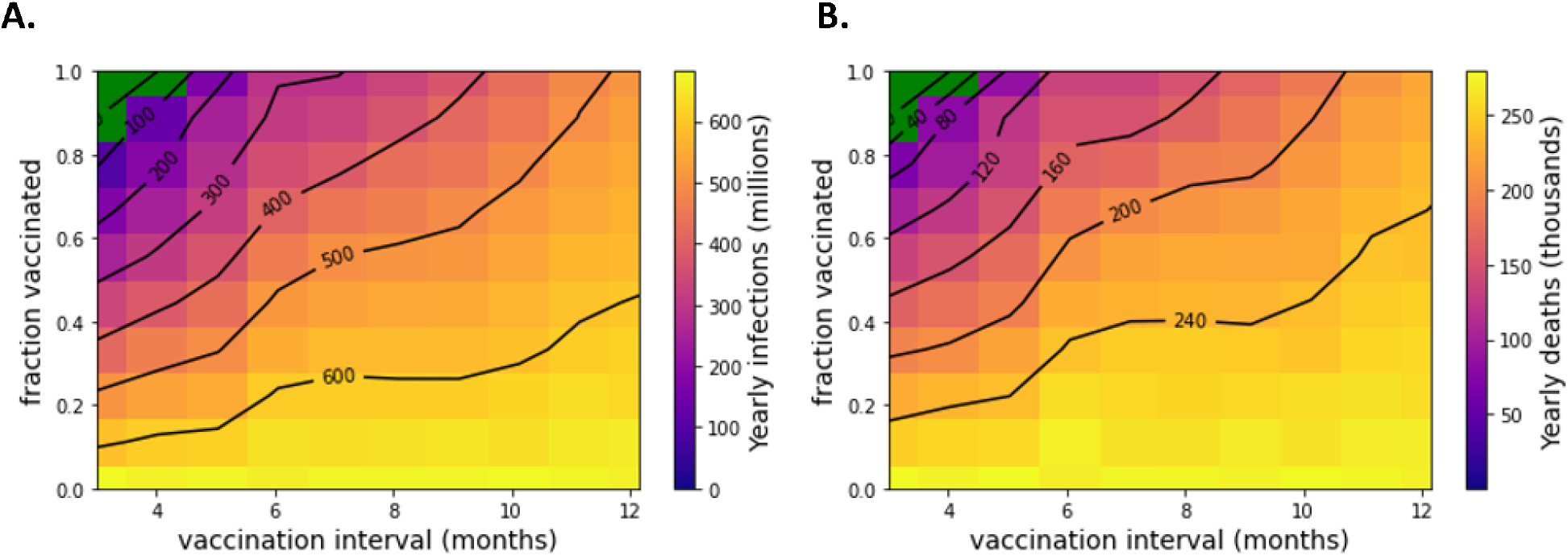
Suppression of SARS-CoV-2 can be achieved with sufficiently high vaccination rates. **A**. Yearly US SARS-CoV-2 infections and **B**. yearly US COVID-19 deaths under a variety of vaccination frequency and compliance scenarios. Green region represents complete suppression (zero infections at steady-state).

### Improved vaccine kinetics improve booster regime efficacy

The Moderna mRNA COVID-19 vaccine induces less durable nAbs relative to post-infection immunity [54,63]. We surmise that, in a future vaccine development program, the target product profile for such a vaccine could be based on ensuring that the distribution of nAb half-lives is improved to match or exceed post-infection immunity.

To evaluate the impact of such improvements, we reimplemented Figure 2 under the assumption that post-vaccination nAbs have the same kinetics as post-infection nAbs (Figure 5). In other work, we found the population median half-life of post-infection nAbs to be 109 days (ref shielding). In this case, the impact of yearly boosters is improved, with lower infection and death rates overall and less variation in outcomes (Figures 5A-C). With a hypothetical vaccine possessing these characteristics, only 3 yearly boosters would be required to achieve near-complete protection from infection in the vaccinated population.

**Figure 5:**
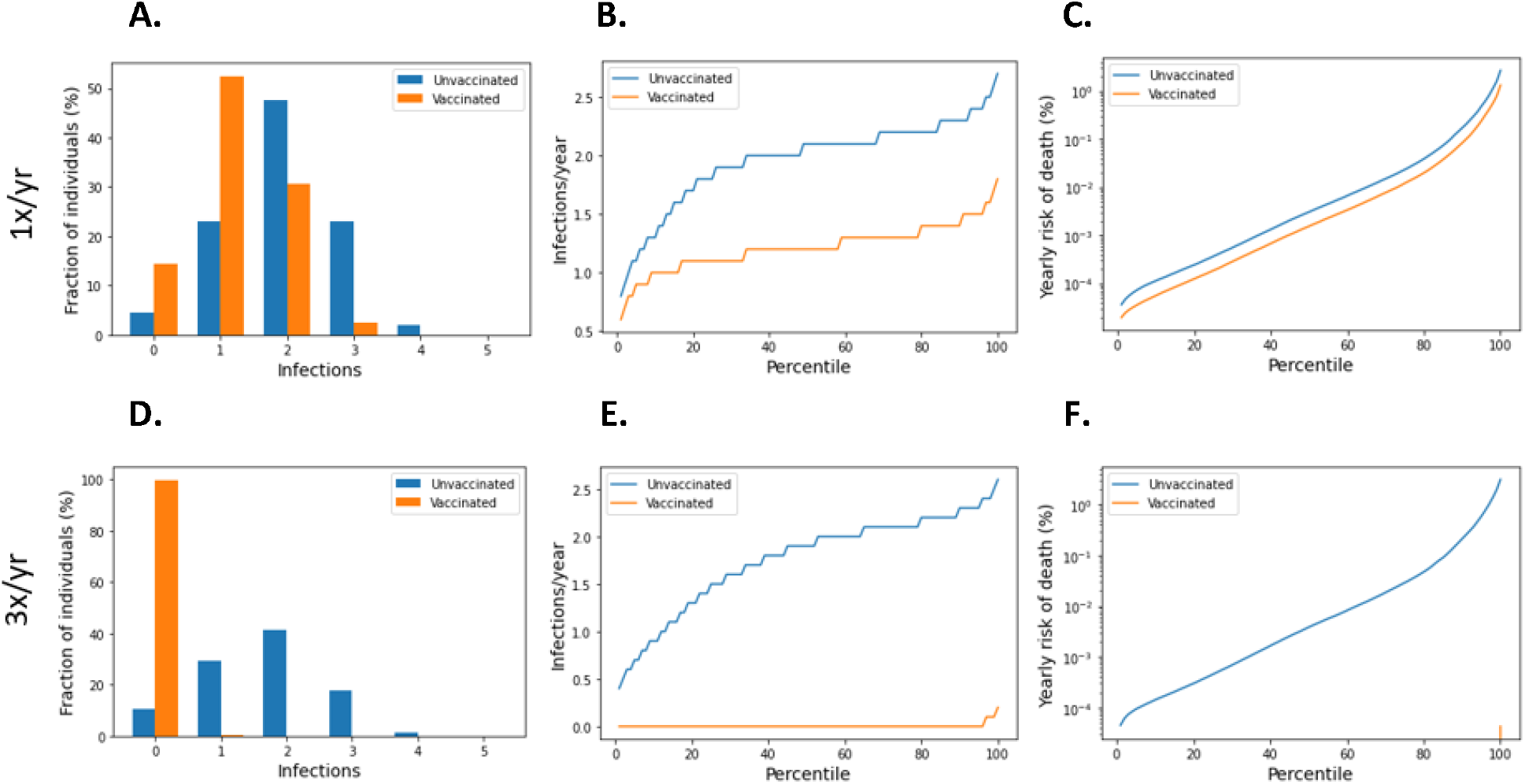
Three yearly boosters may prevent infection in nearly all vaccinees for a vaccine with nAb kinetics similar to post-infection. Column A shows the distribution of infection counts in a single year, which is driven by stochasticity and interindividual heterogeneity. Column B represents the average frequency of infection over a 10-year simulation, at which point interindividual heterogeneity dominates random effects. Column C shows interindividual heterogeneity in yearly risk of COVID-19 death.

### Boosting could have likely averted the delta wave of summer 2021

In Figure 6, we use an SIR model to explore theoretical scenarios similar to the emergence of the delta variant in summer 2021 in the United States. In the early summer, alpha was dominant but in decline, and 48% of the US population was fully vaccinated (see Table 1). As summer progressed, the delta variant established itself in the US [64], leading to a surge in infections. In Figure 6A, we show that unmitigated spread of delta without a booster campaign could have impacted most of the US population (fortunately, infections are estimated to have been limited to approximately 15% of the population [65–67]). Modest (50%) mitigations could have reduced the overall toll, but a large infection count would still be expected (Figure 6B).

**Table 1:** Scenario-specific parameter values for SIRS model.

| Parameter | Value | Units |
| --- | --- | --- |
| Natural immunity decay rate | 0.002 [54] | 1/days |
| Vaccine immunity decay rate | 0.002 [54] | 1/days |
| Delta $R_0$ | 6.0 [103] | Individuals |
| Alpha $R_0$ | 5.3 [104] | Individuals |
| VEi, delta, recent primary series | 70.2 [5] | % |
| VEi, delta, distant primary series | 40.2 [5] | % |
| VEi, delta, recent first booster | 93 [105] | % |
| VEi, pre-delta, primary series | 90 [106] | % |
| VEi, pre-delta, booster | 95 | % |
| Cross-immunity protection | 81 [107,108] | % |
| US adult population | 78 [109] | % |
| US population vaccinated, summer 2021 | 48 [110] | % |
| Transmission mitigation, summer 2021 | 50 | % |

**Figure 6:**
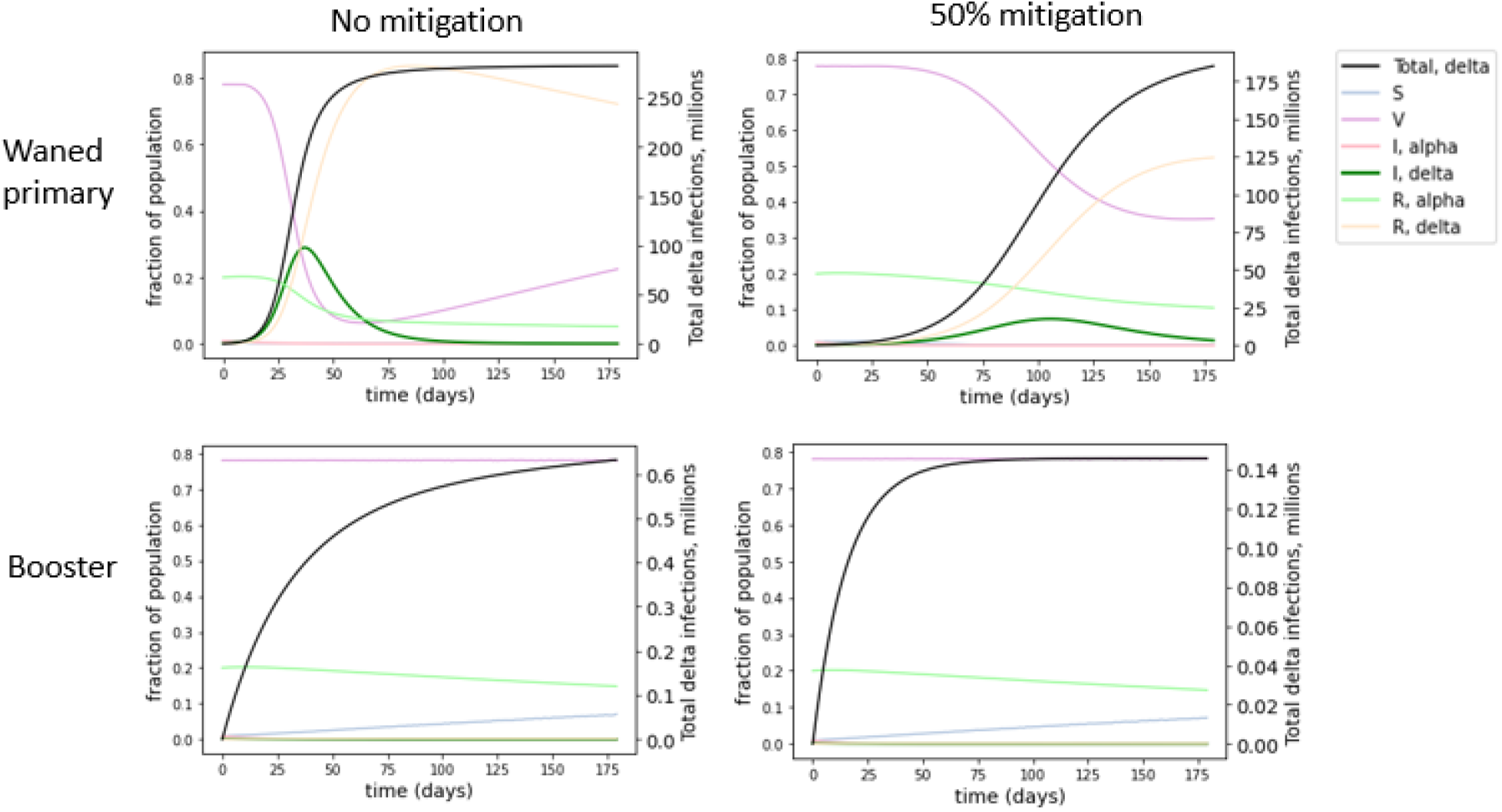
Simulated outcomes for booster roll-out to all US adults under conditions similar to delta emergence. Each compartment in the SIRS model is represented by the fraction of individuals in the population in that compartment over time (left axis). S represents susceptible, V represents vaccinated, I represents infected with alpha or delta, R represents recovered from alpha or delta. The total number of delta infections is tracked (black line, right axis). Regardless of the extent of nonpharmaceutical mitigation of SARS-CoV-2 spread, booster vaccinations for all adults could have significantly reduced delta spread compared to estimated vaccine effectiveness at the time (waned primary series). The SIRS model suggests perfect compliance with a booster campaign before delta became dominant could have averted the delta wave.

However, these outcomes were not inevitable. Even in the absence of NPIs, if all adults had been boosted recently before delta introduction, this could have significantly blunted the impact of the delta surge. Achieving boosting in all US adults could have dramatically improved outcomes, suppressing delta spread to very low levels (Figures 6C, 6D). In Figure S4, we show that boosting only the 48% of Americans who had received the primary series by summer 2021 could have significantly reduced the spread of the delta variant.

## Discussion

The work presented here explores opportunities for improving the performance of the existing SARS-CoV-2 vaccines in mitigating the morbidity and mortality burden of COVID-19. Our findings suggest that vaccine performance may be improved through booster schedule optimization and improved nAb kinetics, providing insights that may be leveraged in the design of further clinical trials. Crucially, our work suggests that boosting 3 or more times a year may preserve VE_s_ and potentially restore VE_i_. Conversely, our findings also suggest that a booster dose frequency of once a year is unlikely to provide a significant population-level benefit from vaccines under endemic SARS-CoV-2 conditions, though individuals may experience transient protection in the post-booster interval.

In this work, we used population PK/PD modeling coupled with an agent-based simulation to interrogate the impact of booster scheduling on vaccinal protection against infection and severe disease. As such, our recommendations provide practical guidance for further vaccine development studies while highlighting the limitations of the current understanding of the immune response to repeated vaccinal dosing. Modeling studies have had substantial predictive value during the course of this pandemic. Our team has used model-based approaches to predict the rapid pace of evolutionary immune evasion [68], the inability of vaccines to enable a return to pre-pandemic conditions [69], the tendency of noncompliance with mitigation measures to incentivize further noncompliance [70], and the rapid variant-driven rebound observed upon premature relaxation of mitigation measures [71]. In each of these cases, our predictions were made months in advance [72–75].

That said, this work has limitations that provide important context for interpreting our findings. First, the extent to which nAbs accumulate upon repeated vaccine dosing is not fully characterized. Some reports have suggested that these antibodies continue to accumulate upon repeated boosting [76–78], while others have suggested a cap or maximum level of neutralizing antibodies [79] or attenuation of response [80]. A recent study conducted by vaccinating Balb/c mice with repeated closely spaced doses of recombinant RBD spike protein reported immune tolerance, albeit with a different dosing schedule and adjuvant from the clinical studies [81], and with the caveats associated with interpreting mouse immunology studies in a human context [82]. Additionally, tolerogenic effects in the study were only observed after the fifth dose, which used a different adjuvant than previous doses, raising the possibility that the change in adjuvant contributed to the change in response [81].

The degree to which homologous boosting with outdated vaccines can raise titers against novel variants is not fully known, nor is the extent to which future nAb responses are shaped by previous exposure (“immune imprinting”, [83]). Despite these concerns, there are reasons for optimism. The bivalent wild-type/BA.5 booster increases neutralizing titers 1.3-fold compared to the monovalent wild-type booster [19], and it increases breadth against emerging immune-evasive variants [84]. Although this model assumes an individual’s nAb vaccinal and post-infection nAb half-lives are fixed, emerging data suggests that nAb half-lives may be increased by repeat exposure [85–88]. Additionally, multiple studies suggest repeat vaccination improves the breadth of the nAb response [86] and antibody avidity [89].

The small population size of the underlying dataset reflects another limitation: the antibody data used to fit the PK model was derived from a Moderna Phase 1 trial enrolling 34 participants, with immunocompromised status being an exclusion criterion for the trial [51,90]. In the model, vaccination status was assumed to be age-independent, and long COVID outcomes were not considered. Finally, the impact of vaccinal side effects or toxicity upon repeated dosing are not considered explicitly, as there is no dataset to define this. It is possible that vaccine toxicity could constrain the frequency of boosting.

As a further limitation, we have not considered the role of compartments of the immune system beyond nAbs in the vaccinal response. Emerging data provides strong support for nAbs as a validated correlate of immune protection (as discussed in the introduction). For further discussion on the role of T-cells in the vaccinal and natural immune response to SARS-CoV-2, see Supplementary Section S1.

To the extent that neutralizing antibodies are the primary correlate of immune protection against SARS-CoV-2, our work makes several crucial points for public-health strategy. First, recommendations for boosting should be driven by science, not based on perceptions of what the public will find acceptable. Current public health messaging is ambiguous around the necessity of booster doses. For example, in the US, public health figures and the administration have expressed a preference for a once-a-year booster focused on the medically vulnerable [49,50,91] and even this has been met with skepticism in some quarters [92,93]. Our work shows that such a strategy would come at a significant human cost, causing VE_s_ to plummet. This messaging has been accompanied by a decline in vaccine uptake, with the monthly averages for US adult vaccines administered having declined ten-fold since its peak in the spring of 2021. At the time of this writing (December 2022), only 22% of US adults have received the most recent booster [65]. A recent Kaiser Family Foundation poll found that 62% of adults are either unvaccinated or are not planning to take the booster at this point. Among people who had already received the original vaccine series but did not intend to receive a booster, the most common reasons for not seeking an additional booster were that they did not believe they needed one (44%) or they did not believe the benefits were worth it (37%) [94]. These numbers are striking given that among individuals who had received the third booster at least four months prior, a fourth booster has been demonstrated to halve infection risk [95] and reduce risk of severe disease by up to 3.5-fold [96].

It is thus imperative that boosting frequency be set by public health authorities on a data-driven basis. If the goal is to implement an “individual public health” strategy, where each person is required to make their own choices to protect themselves from COVID-19, the role of public health in providing honest and accurate feedback around the consequences of those choices cannot be overstated. A thrice-yearly booster frequency may have low uptake, but it may provide better protection for those who seek to avoid SARS-CoV-2 infection.

As a closely related point, our work suggests that repeat boosting is an important tool for maintaining vaccine efficacy between variant-specific vaccine updates, even though new variants often evade existing vaccines to some extent. An additional dose of a partially-matched booster can provide protection against new variants, and this will likely lead to a superior outcome than not taking the dose.

Third, our work suggests the importance of continuing to support alternative dose routes, such as intranasal vaccines, that may possess a more favorable efficacy-to-toxicity ratio (“therapeutic index”) to support more frequent dosing. Exploring alternative dose routes with superior therapeutic indices can also allow for a more complete assessment of alternative boosting frequencies. For example, a weak mucosal immune response from one or two doses of an intranasal vaccine may be compensated for by additional doses if the intranasal vaccine has a higher therapeutic index.

Our work also suggests that nAb persistence is worth optimizing. Hypothetical vaccines inducing more durable nAbs provided better protection and were more likely to lead to elimination of SARS-CoV-2. Future vaccine development work should focus on nAb durability and inter-individual variability as a differentiating factor in the target product profile. This objective may be achievable by changing adjuvants, vaccine formulations, dose, and/or schedule, for example. Preclinical-to-clinical projections of antibody persistence can be used at the discovery stage. Moving into the clinic, the same projections may be used in a Bayesian framework to design parsimonious clinical trials focused on clinical pharmacokinetics to quantify nAb durability in patients and how this varies between individuals. Additionally, interindividual variability in nAb persistence may be shored up by identifying and targeting boosting toward individuals with poor protection (ref).

Finally, our work suggests that suppression of SARS-CoV-2 transmission remains within the realm of possibility. Contrary to perceptions on the topic, improved vaccine design and use may permit suppression of SARS-CoV-2, both for individuals seeking protection from infection and the population as a whole. Repeatedly-dosed vaccines with a longer half-life, a better tolerability profile, or both may allow nAb levels to build up to a point where VE_i_ is maintained even in the face of rapid viral immune evasion. Such a scenario would place suppression of SARS-CoV-2 within reach, particularly if the vaccines are used in conjunction with other measures such as testing, improvements in air quality, and masking.

As we identify a number of immunological open questions, our work suggests that answering these questions should be a focus of research in the near term. In particular, understanding the impact of long-term repeated boosting on nAb production as well as vaccinal side-effect profiles is crucial for enabling more effective use of the existing vaccines. In this context, it bears mentioning that (in the United States as of December 2022) four doses have been recommended for adults in 18 months [97]. Millions of adults in the US have taken vaccine doses on this schedule, and it appears well tolerated. This frequency of vaccine dosing (roughly once every 4 months) is close to the frequency suggested by our work (once every 3 months).

Our work suggests that repeated vaccination is a crucial driver of public health outcomes, but repeated boosting with mRNA vaccines may present tremendous logistical hurdles on a global basis. More readily scaled and distributed technologies (such as spike-protein based vaccines or inactivated vaccines) may also provide acceptable outcomes if patients can tolerate a dosing frequency that is sufficient to enable nAb buildup to the protective levels required to restore VE_s_ (and potentially VE_i_). The side-effect profile of targeted dose routes (such as for intranasal vaccines) should also be examined closely in this context. While manufacturing, tolerability, and compliance constraints may make frequent boosting hard to achieve with the current vaccines, next-generation vaccines should be designed with a target product profile of repeated dosing in mind. For example, room temperature-stable, nasally administered vaccines based on low-cost technologies would make it easier for us to achieve the goal of widespread and repeated vaccinal coverage.

Taken together, our findings suggest that there are ways to improve the utility of the current crop of vaccines in controlling SARS-CoV-2. Our work suggests areas of research focus on open immunological questions that must be resolved in order to establish proof-of-concept for such a strategy. Lastly, our work also suggests a few key properties that the next generation of vaccines should take into account: the durability of nAb response, the tolerability of repeat dosing, and manufacturability at scale. While it is often said that “learning to live with the virus” is inevitable, our work suggests that this is not the case. Improving the performance of existing vaccines and rationally designing future vaccines hold the key to restoring the promise of vaccinal control of SARS-CoV-2.

## Methods

### Agent-based simulation of infection and mortality burden

To determine the impact of boosting regimes on endemic SARS-CoV-2 infections and mortality among the vaccinated and the unvaccinated, we reimplemented an agent-based model developed in our prior work [54]. For this analysis, we updated the model to account for the distribution of neutralizing antibody (nAb) half-lives after vaccination according to our previous mixed-effects model fit (vax heterogeneity paper). Additionally, we extended the model to track individuals’ risk of fatal COVID-19 over time and the total number of COVID-19 deaths in the population.

To account for differences in persistence of post-vaccination and post-infection nAbs, the model separately tracks nAb titers induced by vaccination and infection. Each individual is assigned a vaccine nAb decay rate and an infection nAb decay rate drawn from the respective distributions. Upon vaccination or infection, an individual’s nAb titers are increased by a fixed multiple (revaccination 10-fold [98] and reinfection 14.4-fold [99]). Conservatively, we assumed that any antibodies generated through revaccination (by boosting of pre-existing vaccine or infection-induced antibodies) wane at the vaccine nAb decay rate.

We also updated the model to track the risk of fatal COVID-19. To address interindividual heterogeneity in the risk of severe COVID-19, we introduced age as a property of individuals in the model. Individual ages are set based on a random draw of the age distribution of the United States [100], and the age-related infection fatality rate (IFR) is calculated based on a published formula [101]. We adjusted the population average IFR produced by this model to reflect estimates for omicron’s IFR (0.21%) [102].

Upon successful infection, an individual’s risk of severe disease is also calculated based on their normalized neutralizing antibody titer:

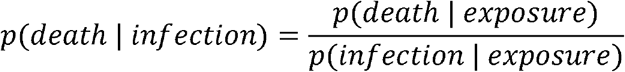

when

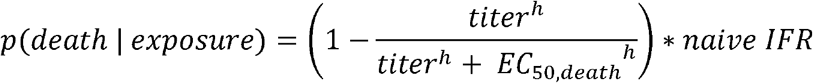

In this case, EC_50,death_ is the neutralizing antibody titer required for 50% protection from death; in this analysis, we assumed EC_50,death_ is equal to the EC_50_ for protection from severe disease parameterized by Davenport et al (3% of CP titer) [33]. The naïve IFR is the individual’s risk of death upon naïve infection. Each individual’s cumulative probability of survival is tracked, with the probability starting at 1 and being multiplied by the risk of death upon each infection. The population-level death count is increased by the expected value of deaths imposed by each infection (that is, the individual’s risk of death for that infection).

In the model, vaccine boosting occurs in vaccinated individuals at a specific revaccination frequency. Each individual’s time since last vaccination is tracked using a counter. At the beginning of the simulation, the counters are randomized over the range 0 to the revaccination interval length (e.g. 180 days for twice-yearly boosting). This ensures boosting in the population is staggered in time, while each individual is revaccinated at exactly the end of their boosting interval.

The model is designed to capture steady-state (endemic) SARS-CoV-2 dynamics. Before each simulation, the model is run for 1000 days to reach equilibrium. To demonstrate both stochastic year-over-year variation in individual outcomes and long-term average risks, we ran simulations over one year and over 10 years after equilibrium conditions were reached.

Thus, this model accounts for inter-individual heterogeneity in nAb waning rates after vaccination [51] and infection [54], a steady rate of nAb potency loss due to immune evasion, variability in contact behavior between individuals, and variation in severe disease susceptibility due to age. For the heterogeneity in outcomes simulations, we assumed that 50% of the population is vaccinated, which is in rough agreement with the fraction of the eligible population that has received a first booster in the US [59]. In the supplement, we explored the outcomes of a negligible vaccinated subpopulation to distinguish first-order (vaccination protects vaccinee) from second-order (vaccination reduces transmission in the population, which protects everyone) effects. We also performed a sensitivity analysis to determine the impact of varying revaccination frequency and acceptance on population-level SARS-CoV-2 infection counts and yearly COVID-19 deaths.

### Susceptible-infectious-recovered-susceptible (SIRS) model of strain invasion

We also performed a retrospective analysis to determine whether a delta-like wave could have been avoided by administering boosters to all US adults under conditions similar to the summer of 2021. For this purpose, we adapted the two-strain SIRS model with vaccines we implemented in our prior work predicting variant-driven waves after vaccine roll-out [71]. We assumed that the alpha variant dominated prior to delta emergence. We assessed multiple scenarios for vaccine efficacy against infection (VEi) based on clinical data for recent two-dose primary series, distant (>90 days) primary series, and recent first booster. We assessed best-case vaccination scenarios in which all US adults were vaccinated and realistic scenarios in which 48% of the population was vaccinated.

## Data Availability

All data produced in the present study are available upon reasonable request to the authors

