## Supplementary Materials for "The impact of vaccination frequency on COVID-19 public health outcomes: A model-based analysis"

^1^ Fractal Therapeutics, Cambridge, MA, USA.

^2^ Dartmouth College, Hanover, NH, USA.

^3^ Stanford University School of Medicine, Stanford, CA, USA.	^4^ Monash University, Melbourne, VIC, AUS.

^5^ Halozyme Therapeutics, San Diego, CA, USA.

^6^ Harvard Medical School, Boston, MA, USA.

^7^ Boston Children’s Hospital, Boston, MA, USA.

^8^ Sage Therapeutics, Cambridge, MA, USA.

^9^ Boston University School of Public Health, Boston, MA, USA.

* Corresponding author

**Abstract:**

While the rapid deployment of SARS-CoV-2 vaccines had a significant impact on the ongoing COVID-19 pandemic, rapid viral immune evasion and waning neutralizing antibody titers have degraded vaccine efficacy. Nevertheless, vaccine manufacturers and public health authorities have a number of levers at their disposal to maximize the benefits of vaccination. Here, we use an agent-based modeling framework coupled with the outputs of a population pharmacokinetic model to examine the impact of boosting frequency and durability of vaccinal response on vaccine efficacy. Our work suggests that repeated dosing at frequent intervals (multiple times a year) may offset the degradation of vaccine efficacy, preserving their utility in managing the ongoing pandemic. Our work relies on assumptions about antibody accumulation and the tolerability of repeated vaccine doses. Given the practical significance of potential improvements in vaccinal utility, clinical research to better understand the effects of repeated vaccination would be highly impactful. These findings are particularly relevant as public health authorities worldwide seek to reduce the frequency of boosters to once a year or less. Our work suggests practical recommendations for vaccine manufacturers and public health authorities and draws attention to the possibility that better outcomes for SARS-CoV-2 public health remain within reach.

**Supplementary file**

**Section S1: Lack of evidence for a protective role for T cell immunity for SARS-CoV-2**

**Summary**

A precise understanding of the role of T cells in COVID-19 is critical for the design of biomedical interventions as well as public health strategy for the ongoing pandemic. Here we discuss the relative importance of T cells in protecting against infection by the SARS-CoV-2 virus and also against the risk of developing severe COVID-19. We will contrast this role against the role played by neutralizing antibodies (nAbs) produced as part of the B cell-mediated (humoral) adaptive immune response.

At this point, the weight of evidence argues against a role for T cells in providing protective immunity against SARS-CoV-2. Early in the pandemic, a body of studies demonstrated convincingly that the T cell response is durable after both infection and vaccination (See **Footnote A** below for details). These encouraging findings led to the widely held perception that sustained immunity against severe disease [1–3] and protection against viral immune evasion [4, 5] would be provided by the observed durability of T cell responses. This hope was widely expressed in the public sphere as well [1, 2, 6, 7]. However, vaccinal immunity against severe disease and death (VE_s_) has been observed to wane rapidly over time and in response to viral evolution (See **Footnote B** below). This rapidly waning VE_s_, despite the presence of a durable T cell response, creates a logical contradiction in the proposition that T cells play a role in providing protective immunity.

In addition, while certain pools of T cells (in particular SARS-CoV-2-specific CD8+ and CD4+ T cells) are frequently found in convalescent patients, overall T cell numbers decrease during infection with SARS-CoV-2, in both mild and severe cases (See **Footnote C**), with T cells being infected directly by the virus and undergoing apoptosis as a result (**Footnote D**). Conflating the post-infection increase in the relative frequencies of CD4+ and CD8+ T cells with their absolute numbers is a source of confusion on this point. Further, the loss of T cells (either due to disease or drug treatment) does not result in worse outcomes for COVID-19 disease progression (See **Footnote E**), and the T cell blockading drug Abatacept has shown a positive impact on disease outcome in COVID-19 patients in a clinical setting (**Footnote E**), suggesting a net negative impact of T-cell response on disease pathophysiology. Notably, SARS-CoV-2-specific CD8+ and CD4+ T cells have also been found in unexposed controls from the beginning of the pandemic; these are thought to arise as a result of cross-reactivity with common cold coronaviruses (**Footnote C**). Despite reports to the contrary, T cells levels are not predictive of protection against SARS-CoV-2 infection (**Footnote F**). Similarly, reports of T cell-mediated protection in the absence of a functional B cell (or nAb) response have been prone to issues with interpretation of the underlying data (**Footnote G**).

On the other hand, a strong case can be made that protection against SARS-CoV-2 infection is provided by nAbs. nAbs are a validated correlate of immune protection, and they have been shown in multiple studies to be predictive of vaccinal protection for both symptomatic infections and severe disease, across a range of vaccines and viral variants (See **Footnote H** for details). Drugs and conditions that impair B cell function or lead to lower nAb levels are strongly associated with worse outcomes for SARS-CoV-2 infection (see **Footnote I**). Restoration of nAb levels is strongly associated with improved outcomes (See **Footnote J**). Limited preclinical data suggests that this is not the case for T cell functionality for SARS-CoV-2 (**Footnote J**).

The observed evolutionary trajectory of the SARS-CoV-2 virus adds further support for the argument that humoral and not cellular immunity is the primary source of protection for COVID-19. It is a basic principle in evolutionary biology that the strength of the selection pressure determines the rate of evolution of resistance [8, 9]. Currently, viral evolution is being primarily driven by immune evasion- specifically against nAbs [10, 11]. We predicted this rapid evolution of immune evasion using an evolutionary biology framework in a paper [12] whose preprint was posted in the fall of 2020 [13]. That work started from the premise that neutralizing antibodies would exert a selection pressure that the virus would need to evade, and we demonstrated that rapid immune evasion could be possible under conditions where there was a large standing pool of SARS-CoV-2 infections. Thus, rapid evolution of immune evasion to nAbs validates their impact on viral fitness. Conversely, the evolutionary stability of the T cell epitopes is consistent with no selection pressure being exerted by the T cells on SARS-CoV-2. (The alternative explanation for the evolutionary stability of the T cell epitopes, that they cannot be evaded by the virus, has been undermined by the rapid reductions in VE_s_ due to evolutionary immune evasion).

In summary, multiple lines of evidence argue against a role for T cells in protection against SARS-CoV-2 infection. The observed durability and evolutionary stability of the T cell response has not translated into lasting protection against infection or severe disease.

**Footnotes:**

**A: T cell response is persistent and evolutionarily stable**

T cell response is durable after infection [14–17]. The fraction of spike-specific T cells goes up rapidly upon vaccination [18], and the T cell response in vaccinated individuals is also persistent [1, 2, 14]. T cell (but not B cell) epitopes are evolutionarily stable- while B cell epitopes in the spike glycoprotein (S) and in the nucleocapsid protein (N) have higher diversity than nonepitope positions, epitopes for CD4+ and CD8+ T cells are not more variable than nonepitope positions [19]. The T cell response remains durable even in the face of the newer immune-evading variants [5, 20–24] and the vaccinal and natural T cell responses to Omicron BA.1, for example, are broadly preserved [25–27].

While a number of papers have reported immune escape mutations at T cell epitopes, we note that many of these mutations are not present in the currently circulating SARS-CoV-2 lineages (Table 1)

| Protein | Substitution | T cell class | Current frequency | Reference |
| --- | --- | --- | --- | --- |
| S | N969K | CD4+ | 93% | [28] |
| S | P272L | CD8+ | 0% | [29] |
| S | A1022S | CD4+ | 0% | [30] |
| S | T1072I | CD4+ | 0% | [30] |
| S | E484K | CD8+ | 0% | [31] |
| N | T362I | CD8+ | 0% | [32] |
| N | P365S | CD8+ | 0% | [32] |
| N | P13L | CD8+ | 93% | [32] |
| ORF3a | Q213K | CD8+ | 0% | [32] |
| ORF1a | T147I | CD8+ | 0% | [33] |
| N | P344L | CD8+ | 0% | [33] |
| M | A69V | CD8+ | 0% | [33] |
| ORF1a | I2230T | CD8+ | 0% | [34] |
| S | L452R | CD8+ | 73% | [34] |
| S | L822P | CD8+ | 0% | [33] |

Table 1: Reported immune escape mutations in the T cell epitopes of the spike protein, with their current frequency in the cov-spectrum database (as of 01/19/23) [35]. List is not exhaustive, see [36] for additional examples of reported T cell-evasive mutations.

**B: Vaccinal efficacy against severe disease (VE_s_) is limited, and vulnerable to immune evasion.**

)A recent CDC study showed that the vaccine effectiveness (a measure of VE_i_) of a bivalent mRNA COVID-19 booster received after 2 or more doses of monovalent vaccines ranged from 43% (for the 18-49 age group) to 22% (for the over-65 age group) [37]. In the original placebo controlled trials among mostly naïve individuals, absolute vaccine efficacy (VE_s_) was originally observed to be very high [38, 39], however over time this efficacy waned in concert with nAb titers in both placebo-controlled trials [40] and large-scale observational studies before the omicron wave dramatically increased the background rate of immunity [41–45]. More recent trials in much more extensively vaccinated and infected populations confirm declines in VE_s_ over time that can be increased by boosting [46]. When it comes to severe acute disease, VE_s_ for a newly boosted individual is now 56% [40]. Continued viral evolution has been degrading VE_s_ [47–50], although it is partially restored with boosters [51]. Two-dose vaccinal series have only a modest benefit (VE_s_~50%) against severe disease with Omicron [47, 52–54] despite these vaccinations having been demonstrated to elicit a T cell response that is both persistent and conserved across variants of concern [21–23, 52].

**C: Issues with interpretation of T cell levels**

Both unexposed individuals and those convalescing from severe COVID-19 have SARS-CoV-2-reactive T cells [55–57], particularly CD4+ and CD8+. Some proportion of these SARS-CoV-2 reactive T cell populations are cross-reactive with common cold coronaviruses [56, 58–60]. While SARS-CoV-2-specific CD8+ and CD4+ T cells are more commonly found in convalescent patients than in unexposed controls, overall T cell levels are not elevated in convalescent patients [56]. Notably, the absolute counts of total lymphocytes and CD4+ and CD8+ T cells in hospitalized as well as non-hospitalized COVID cases are lower than those in healthy counterparts ([61], see Table 1 and 2; [62], see Table 3; [63], see Figure 1; [64], see Figures 1 &2 ).

**D: SARS-CoV-2 infection actively suppresses the T cell response.**

SARS-CoV-2 inhibits the induction of the MHC-I pathway in cells infected by the virus (thereby evading T cell recognition) by several different molecular mechanisms [33, 65, 66]. Viral infections also impair the capacity of monocytes to activate SARS-CoV-2 CD4+ and CD8+ T cells [67]. In addition to impairing T cell activation, SARS-CoV-2 also damages and depletes T cells directly [68]. T cells are directly infected by SARS-CoV-2 [69, 70], express markers of functional exhaustion following severe COVID [71, 72] and undergo frank apoptosis during viral infection [71, 73, 74]. Notably, T cell lymphopenia and dysregulation are observed in both mild (non-hospitalized) and severe cases of COVID [61, 71–77].

**E. Loss of T cell function does not negatively impact outcomes upon SARS-CoV-2 infection.**

T cell depletion via disease or therapeutic intervention has not directly been demonstrated to lead to worse outcomes for SARS-CoV-2 infection. A meta-analysis of outcomes of patients with HIV and SARS-CoV-2 coinfection showed that HIV was not associated with the severity of the disease (OR: 1.28; 95% CI 0.77-2.13, 13 studies), or death (OR: 0.81; 95% CI 0.47; 1.41, 23 studies) in COVID-19 patients, although HIV was independently associated with an increased risk of hospitalization (OR: 1.49; 95% CI 1.01-2.21, 6 studies) and death (hazard ratio: 1.76, 95% CI 1.31-2.35, 2 studies) compared to HIV-negative individuals [78]. Direct depletion of T cells similarly has no impact on COVID-19 outcomes. T cell depletion in macaques does not lead to higher viral loads in the lung [79], nor does it disrupt protection against short-term reinfection [80]. Similar findings have been reported with the ablation of T cell function in humans. Abatacept, used for treatment of rheumatoid arthritis (RA), blocks T cell co-stimulation and decreases T cell activation [81–83]. Abatacept treatment is not associated with worse outcomes for RA patients infected with SARS-CoV-2 (HR: 1.14, 95% C.I.: 0.77 to 1.68), while B cell (humoral immunity)-depleting therapies lead to significantly worse outcomes in the same population: Rituximab (HR: 4.25, 95% C.I.: 3.17 to 5.69) and Janus Kinase inhibitors (HR: 2.05, 95% C.I.: 1.57 to 2.69) [84]. In fact, Abatacept treatment improves outcomes for COVID-19 [85, 86], consistent with the negative role for T cells in the disease that has been proposed by a number of researchers [87–90].

**F. T cells are not predictive of protection against SARS-CoV-2 infection.**

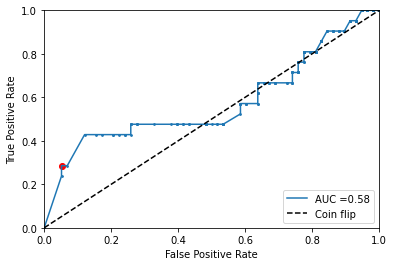
The presence of SARS-CoV-2- specific T cells does not predict for protection against SARS-CoV-2 infection. We analyzed the dataset from Dowell, et al. [91] (examining the frequency of spike-specific T cells and subsequent positive SARS-CoV-2 test results in children) using a receiver operating characteristic (ROC) analysis [Fig S1]. Although the authors suggest an association between the presence of T cells and protection against infection, we found that this association is weak. In this study, the authors measured spike-specific T cells and anti-spike IgG titers in enrollee children. The children’s health records were monitored for later confirmed COVID-19. The authors selected thresholds for cellular and antibody positivity and assessed these as predictors of later test positivity. However, threshold choice can be expected to impact the apparent predictiveness of cellular and antibody positivity [92]. To assess the general predictive value of spike-specific T cell frequency, we constructed an ROC curve using the data in the study. We found that the area under the ROC curve (ROC AUC) is low at 0.58. For reference, a purely random predictor (i.e., a coin flip) will have an ROC AUC of 0.5, while a perfect predictor will achieve an ROC AUC of 1.

Figure S1: ROC curve describing spike-specific T cell frequency as a predictor of testing positive for SARS-CoV-2. Red point represents threshold selected in the study.

**G. Studies reporting T cell protection in the absence of nAbs are prone to interpretation challenges.**

Both antibody and T cell responses are induced by primary SARS-CoV-2 infection in unvaccinated individuals, and the strength of the response across both arms of the immune system is correlated [56]. This makes it challenging to interpret studies attributing outcomes specifically to T cell function. For example, a recent study claimed that vaccines eliciting a stronger T cell response provided protection against SARS-CoV-2 infection in a murine model [93]. However, the vaccines that elicited higher levels of CD4+ T cells also elicited higher levels of nAb response, complicating the attribution of the improved protection to CD4+ T cells (See fig 1A and B in [93]).

Similar challenges have been associated with clinical studies- a number of studies have attempted to attribute the increased severity of COVID-19 in cancer patients, for example, to impaired T cell functionality [94, 95]; however, these patients often have an impaired nAb response as well [95–97]. A recent study identified a subset of patients with defective humoral immunity whose survival they attributed to CD8+ T cell counts ([95], see figure 4H). However, in this high-dimensional dataset, they selected a threshold for CD8 counts using a Classification and Regression Tree (CART) analysis, without specifying the use of a separate test and training set. The lack of cross-validation for a selected threshold makes the result challenging to interpret (as discussed in **Footnote F** above)- the result obtained here had a p-value of 0.04, which was not corrected for multiple comparisons. The lack of multiple comparisons corrections for statistical analysis on high-dimensional datasets is seen in other papers that claim an association between T cells and protection from infection more generally. For example, a recent paper [58] claimed that cross-reactive T cells protect against SARS-CoV-2 infection. The argument put forth by the authors hinges on a difference in frequency of IL2-secreting (but not IFN-γ or dual-secreting) cross-reactive T cells, with an uncorrected t-test p-value of 0.0139. (See Figure 2. Notably, this was one of 12 comparisons shown in the figure!).

In other cases, study conclusions directly differ from the observed data. For example, one study claiming a role for CD8+ T cells in protection against SARS-CoV-2 showed that CD8+ immunodepleted macaques had no detectable viral load in the lungs upon viral rechallenge (similar to control, [79], see fig 4).

**H. nAbs are a correlate of immunity**

Neutralizing antibody (nAb) titers are a validated correlate of immune protection [53, 98–104] for SARS-CoV-2, as they are for other viruses [105]. nAb titers normalized to mean convalescent titer (from the same study) have been shown to fit well to a nonlinear dose-response relationship that is predictive of reported vaccinal protection across a range of different vaccines [101]. Two such dose-response curves exist, one linking nAb titers to protection against symptomatic infection, and one linking nAb titers to protection against severe COVID-19 outcomes. These relationships have held up across a range of studies [102, 103], and are also predictive of vaccinal efficacy against newly emerging variants [51, 104, 106–108]. Neutralizing antibody titers correlate with waning vaccinal protection against infection (VEi) due to PK waning [102, 109] or viral immune evasion [51, 104, 106–108] and with loss of vaccinal efficacy against severe disease (VEs) [101].

**I. Loss of B cells or nAbs leads to dramatically worse outcomes upon SARS-CoV-2 infection**

Rituximab suppresses the B cell, but not the T cell response for COVID-19 [110], and has been reported to lead to an increased risk of severe disease and/or death for COVID-19 patients in several studies [84, 111–115]. Similarly, CD20 therapy for multiple sclerosis, which suppresses humoral, but not cellular responses [116, 117], has been associated with an elevated risk of severe disease or death due to COVID-19 [115, 118]. Of note, these findings extend to vaccinal protection as well. Immunosuppressed patients have an impaired humoral, but not cellular response to COVID-19 vaccination [119]. Breakthrough infections for immunosuppressed patients have poor outcomes [120, 121], and VE_s_ in these populations is lower than that of the immunocompetent population [122–124].

**J. Restoration of B cell (antibody), but not T cell function, mitigates COVID-19 severity**

Prior to the evolution of viral resistance, monoclonal antibodies had a strongly beneficial impact on COVID-19 disease outcomes [125]. Conversely, B cell depleting therapies lead to worse outcomes for breakthrough infections [120], which during the early phase of the pandemic could also be rescued by the use of monoclonal antibody therapies [120]. Vaccines that specifically generate a T cell response have minimal impact on COVID-19 in mouse models, while those that generate a B cell response do [126]. Robust T cell responses are not associated with recovery in critical COVID-19 patients [127]. Crucially, T cell levels take months to return to normal ranges after severe COVID [56, 128], which is inconsistent with the strong protection against reinfection typically observed immediately after recovery from acute COVID [129, 130]. (A subset of patients with mild-to-moderate SARS-CoV-2 infections also experience T cell depletion (including the loss of naïve T cells) and dysregulation [77].

**Acknowledgements**

The authors acknowledge Ms. Gerda Lobo for critical reading and feedback on the manuscript.

**Supplementary Figures**

**A. B.**

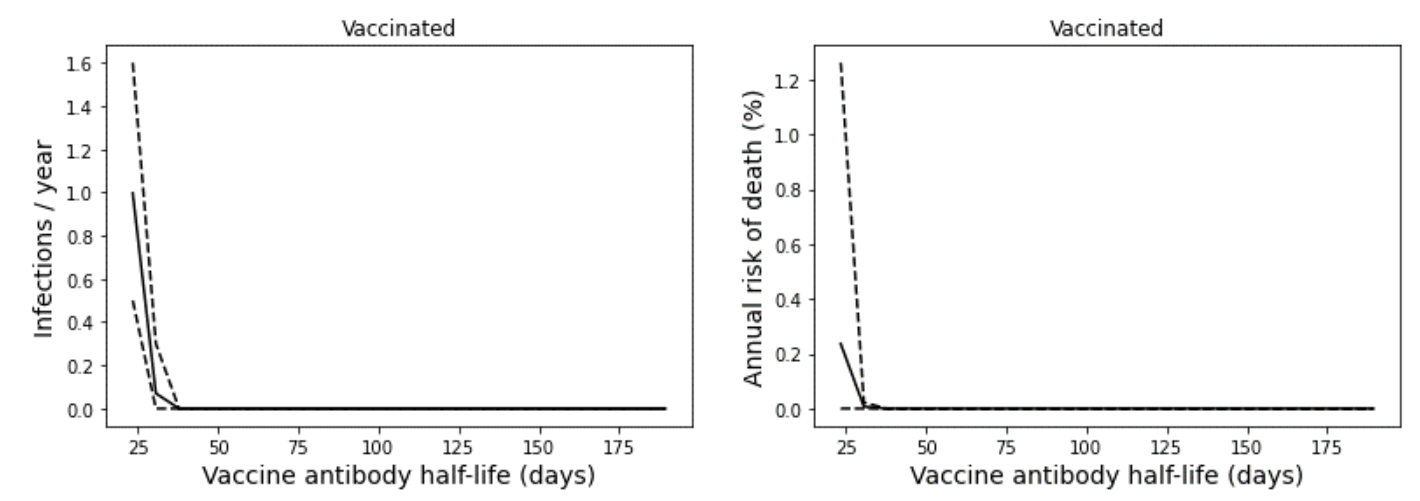

**Figure S2:** A more frequent vaccination schedule using current vaccines may benefit individuals with the shortest vaccine nAb half-lives. In this example, a five times yearly booster schedule is shown to prevent infections in all individuals with at least 40-day vaccine nAb half-lives. A six times yearly schedule was found to prevent infections in all immunocompetent individuals above the first percentile for vaccine nAb half-life. **A.** Average infection frequency and **B.** average yearly risk of death over a 10-year simulation. Dashed lines represent the 90% population interval.

**A. B.**

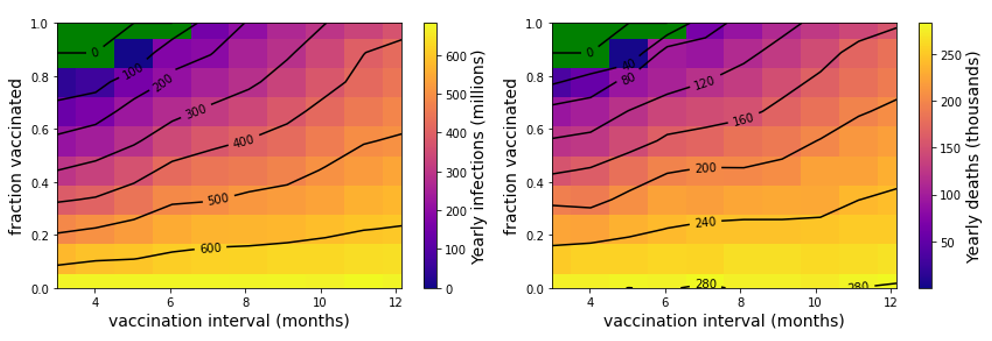

**Figure S3.** Increased durability of vaccine nAb titers improves population-level COVID-19 outcomes. **A.** Yearly US SARS-CoV-2 infections and **B.** deaths are simulated under a variety of vaccination frequency and compliance conditions. Green region represents complete suppression of SARS-CoV-2 transmission.

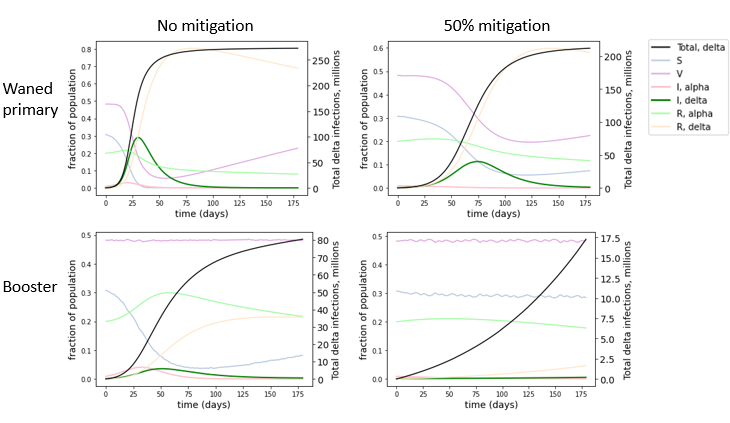

**Figure S4:** Boosters in the summer of 2021 could have blunted the impact of the delta wave in the US, even if greater adult compliance could not be achieved. In this figure series, we assume 48% of the population is boosted; this is the fraction of the US population that had received the primary series at the time [110].

A. **B. C.**

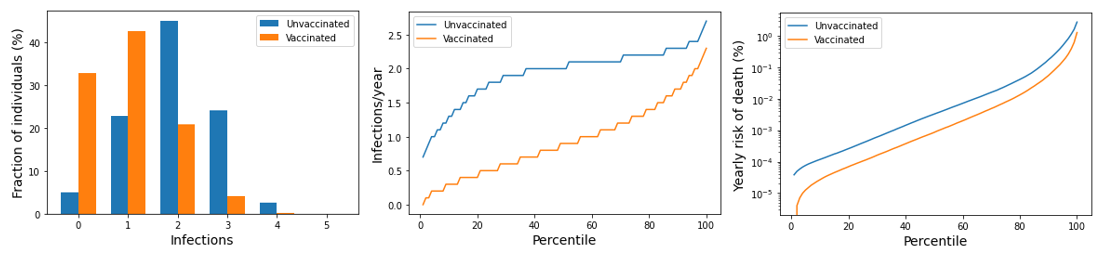

**D. E. F.**

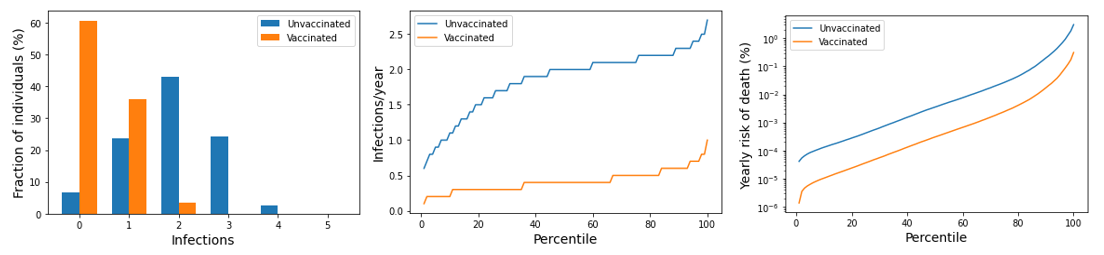

**Figure S5.** Impact of twice-yearly boosters on COVID-19 outcomes. Impact of simulated Moderna boosters on **A)** distribution of total infections in a given year, **B)** average yearly frequency of infection and **C)** average yearly risk of COVID-19 death. Impact of boosters with post-infection-like nAb kinetics on **D)** distribution of total infections in a given year, **E)** average yearly frequency of infection and **F)** average yearly risk of COVID-19 death.

A. **B. C.**

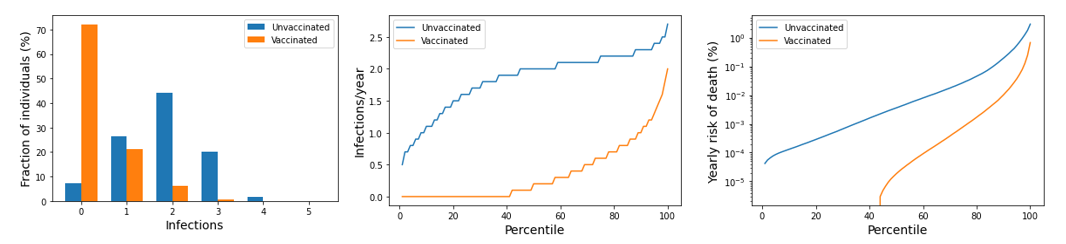

**Figure S6.** Impact of thrice-yearly Moderna boosters on COVID-19 outcomes. Impact of simulated Moderna boosters on **A)** distribution of total infections in a given year, **B)** average yearly frequency of infection and **C)** average yearly risk of COVID-19 death.
